# Integrating AI-ECG and Point-of-Care Cardiac Ultrasound for Screening Structural Heart Disease: A Proof-of-Concept Study

**DOI:** 10.1101/2025.04.24.25326391

**Authors:** Francisco B. Alexandrino, Reid Schlesinger, Jared Bird, Eunjung Lee, Abhishek J. Deshmukh, Vuyisile T. Nkomo, Jae K. Oh, Peter A. Noseworthy, Patricia A. Pellikka, Zachi Attia, Francisco Lopez-Jimenez, Paul A. Friedman, Garvan C. Kane, Sorin V. Pislaru, Gal Tsaban

## Abstract

**Background:** Early structural heart disease (SHD) detection is crucial for improving prognostic outcomes, but widely accessible screening methods are lacking. The advent of artificial intelligence-enabled electrocardiograms (AI-ECG) and point-of-care cardiac-ultrasonography (POCCUS) offers promising new approaches for patient screening. We explored the feasibility and potential of integrating these innovative technologies into a practical SHD screening framework.

**Methods:** Outpatients who underwent ECG at the Mayo Clinic electrocardiogram-laboratory between November 2023 and February 2024 were randomly offered also to receive POCCUS, performed by a novice operator and reviewed by an expert echocardiologist. Impressions from AI-ECG and POCCUS were integrated to assess for SHD, including low left ventricular systolic function (ejection-fraction<50%), aortic stenosis, and increased left ventricular wall thickness indicative of cardiac amyloidosis or hypertrophic cardiomyopathy. Operators were blinded to patients’ comorbidities and formal echocardiogram results.

**Results:** Of 486 patients (median-age 64 years;49% women), 286 had available formal echocardiography, with 17.5% having SHD. AI-ECG had a 32% positive predictive value (PPV) and a 94% negative predictive value (NPV) to detect any SHD. Adding POCCUS increased the overall PPV to 64% with an NPV of 93%, with an increase in diagnostic accuracy from 67% to 88%. Notably, 89.5% (17/19) of the “false positives” by AI-ECG+POCCUS had less-than-moderate-SHD. Applying the AI-ECG+POCCUS screening workflow on the entire cohort resulted in a number-needed-to-screen of eight to identify one patient requiring formal echocardiography (Central Figure).

**Conclusions:** The integration of AI-ECG and POCCUS holds promise as a potentially effective screening method for SHD, facilitating improved patient selection for formal echocardiography.

## INTRODUCTION

Structural heart diseases (SHD), including left ventricular systolic dysfunction (LVSD), aortic stenosis (AS), cardiac amyloidosis (CA), and hypertrophic cardiomyopathy (HCM), pose significant risks of heart failure and mortality, often evading detection until advanced stages or fatal outcomes manifest [1–4]. Advances in medicine have introduced treatments that alter the course of SHD, emphasizing the critical need for early diagnosis and proactive clinical and imaging follow-up to identify candidates for beneficial interventions [5,6]. Thus, standardized, easy-to-implement, and cost-effective screening measures for SHD are needed.

Artificial intelligence-enabled electrocardiogram (AI-ECG) algorithms have proven effective in detecting SHD in the general population, boasting high accuracy with areas under the curve ranging from 0.85 to 0.96 and an impressive negative predictive value (NPV) nearing 99% [7–10]. However, their low positive predictive value (PPV; 11%-34%) limits their utility as standalone screening tools, potentially leading to frequent unnecessary and costly transthoracic echocardiograms (TTEs). This issue is compounded by the overuse of expensive diagnostics and the constrained capacity of echocardiography laboratories to manage screening TTEs for patients flagged by positive AI-ECG results, necessitating an innovative approach.

Recent advances in medical technology have facilitated the development of compact, high-quality portable ultrasound devices. The emergence of small, handheld ultrasound machines capable of acquiring high-resolution 2D echocardiography images has transformed point-of-care assessment. Portable, cost-effective, and increasingly deployed across various clinical settings, point-of-care cardiac-ultrasonography (POCCUS) enables straightforward qualitative evaluation of cardiac function and structure through simple two-dimensional imaging [11].

Considering these developments, integrating POCCUS screening with current AI-ECG models presents a promising and underexplored opportunity to identify patients with SHD. Outpatient ECG labs, bustling with activity, are ideal settings where AI-driven ECG analyses are routinely conducted. This environment offers a practical, real-world context for combining ECG and POCCUS for screening purposes. The current study aims to assess the feasibility and predictive implications of integrating AI-ECG and POCCUS screening for SHD within a diverse outpatient population.

## METHODS

### Study design

We conducted a proof-of-concept quality-of-care project at the ECG Laboratory of Mayo Clinic in Rochester, Minnesota, USA. This analysis aimed to assess the feasibility and diagnostic yield of a unique screening framework employing a two-layer screening process integrating AI-ECG and POCUS classifications.

To explore the feasibility of integrating POCCUS scanning into the ECG lab, the Mayo Clinic Echocardiography and ECG laboratories initiated an in-house quality-of-care project. Novice physician operators with no echocardiography experience (FBA, RS) were trained in-house to acquire POCCUS imaging was present in the ECG lab approximately three days a week between November 1, 2023, and February 28, 2024. Adult (≥18 years old) all-comers undergoing ambulatory ECG on days when POCCUS operators were available were offered a POCCUS screen after their ECG. Interested patients were briefed by the ECG technician and the POCCUS operator and provided oral consent before scanning. The POCCUS studies were uploaded and stored on an encrypted online patient-care platform after acquisition and subsequently reviewed by a trained cardiologist with 8 years of experience (GT) to assess the likelihood of structural heart disease (SHD). Any abnormal findings not previously documented were communicated to the patient’s primary provider. Patients with poor-quality POCCUS images were excluded. No further exclusion criteria were applied. The Mayo Clinic institutional review board approved the project.

### AI-ECG

The Mayo Clinic ECG Laboratory performs an average of 250 ECGs daily. All ECGs are acquired by a technician in the supine position at a sampling rate of 500 Hz using a GE-Marquette ECG machine (Marquette, WI). The data are stored using the MUSE data management system (GE Healthcare, Chicago, IL) for review and immediate clinical interpretation. AI-ECG data, generated through validated algorithms, becomes accessible upon confirmation of the ECG clinical interpretation report. This data provides probabilities and categorizations for aortic stenosis, decreased left ventricular ejection fraction (LVEF), HCM, and CA using specific thresholds (40.6%, 25.6%, 11%, and 48.5%, respectively. These data are available for healthcare providers to review on a separate platform linked to the patient’s Electronic Medical Record. Of note, the cut-off for decreased LVEF in the AI-ECG algorithm used in the current study was validated for detecting LVEF <35%. However, its application in the general population also demonstrated an AUC of 0.880 for identifying LVEF <50% [12].

### POCCUS

Novice POCCUS operators, first-year postgraduate physicians with no prior cardiac ultrasound training, participated in a structured, two-day training program led by an expert advanced cardiac sonographer. POCCUS was conducted using a portable Lumify^™^ S4-1 handheld ultrasound device (Phillips Healthcare). Lumify^™^ S4-1, as most hand-held devices, doesn’t have quantitative Doppler. Patients were imaged in the supine position. Clips were acquired in 10-second loops from parasternal (long and short axes), apical (four chambers), and subcostal acoustic windows. The studies were saved with patient-identifier numbers, uploaded, and stored on a dedicated, secured POCUS review platform (Qpath, Telexy, Canada). Subsequently, a trained cardiologist reviewed the studies to assess the likely presence of SHD. Interpretation relied on visual assessment of the two-dimensional POCCUS studies. Suspected LVSD was defined as LVEF less than 50%, possible AS was defined by significant calcification or restricted excursion of the aortic valve leaflets, and increased left ventricular wall thickness (ILVWT) was determined by offline assessment of LV walls. The physician operators conducting POCCUS and the interpreting cardiologists were blinded to patients’ medical records and AI-ECG results. The quality of the ultrasound clips obtained by the novice sonographers was rated on a scale from 1 to 4, with 1 indicating the highest quality and 4 representing low quality. The average quality scores for each acoustic window were as follows: parasternal long axis (2.5), parasternal short axis (2.6), apical (2.7), and subcostal (2.5).

### Data Collection

All baseline characteristics, encompassing demographics, medical comorbidities, cardiovascular history, and results of formal echocardiography studies, were extracted and validated through meticulous manual chart review.

### Statistical analysis

The main aim of the current analysis was to examine the feasibility and diagnostic yield of incorporating AI-ECG and POCCUS to screen for SHD in a nonselective patient population. Since operators were blinded to the patient’s medical history and diagnostic workup, the study population comprised patients with and without recently available formal TTE evaluation. 121 (42%) of TTEs were done on the same day as the POCCUS and only 39 (13%) were done ≥ 1year before the POCCUS. Therefore, the analysis was done in two stages. First, we examined the NPV and PPV of AI-ECG only and AI-ECG and POCCUS in identifying patients with verified SHD by TTE (*Supplement 1*). In the next stage, we incorporated data among all study participants to assess the number needed to screen according to a proposed two-layered screening method utilizing AI-ECG followed by POCCUS. In the first stage of our analysis, we assessed the PPV and NPV of AI-ECG in comparison to the TTE. In the second stage, we utilized the classifications derived from both AI-ECG and POCCUS to determine which patients required referral for formal TTE evaluation. Patients with previous Aortic Valve Replacement (AVR) were excluded from the diagnostic yield calculation to detect AS; however, they were included in the analysis for LVD and WT. Data are presented as means and standard deviations or median and interquartile range (IQR) for continuous variables, per the variable’s distribution, and as frequencies and percentages for categorical variables. Comparisons between groups were performed using chi-square tests for categorical variables and independent T-tests or Mann-Whitney U test for continuous variables, according to the variable’s distribution.

Baseline characteristics of the study population are presented across the availability status of formal TTE evaluation. We assessed the NPV and PPV values of AI-ECG only and AI-ECG combined with POCCUS vs. formal TTE evaluation (referred to as the gold standard). Next, we simulated a potential workflow matrix where ECG is first implemented to screen out patients “negative” for SHD, followed by a second layer of POCCUS screening for patients deemed as “positive” for any SHD by AI-ECG. According to this suggested workflow, only patients deemed as “positive” for any SHD by both modalities would be defined as likely to have clinically relevant SHD and would qualify/prioritized for formal screening with TTE. The number needed to screen to detect one patient eligible for formal evaluation was calculated for any SHD and individual SHDs based on the rates of “double positive” patients out of all screened patients. A two-sided p-value of less than 0.05 was considered statistically significant for all analyses. Statistical analysis was performed using the JMP software version 17.0.0.

## RESULTS

During the days of available POCCUS imaging in the ECG lab, an average of 250+32 ECGs were performed in the ECG lab every day. A daily average of 25±7 POCCUS studies were performed, corresponding to 10-15% of all daily ECGs. The average duration of the POCCUS studies was 6±2 min from when the ECG was done until the POCCUS operator left the room.

A total of 500 patients were included in the study. A total of 14 patients were excluded because of unavailable POCCUS studies: eleven due to upload and storage failures and three due to uninterpretable imaging. This resulted in a final study population of 486 patients, 286 of which with a previous formal echocardiogram (*Figure 1*).

**Figure 1:**
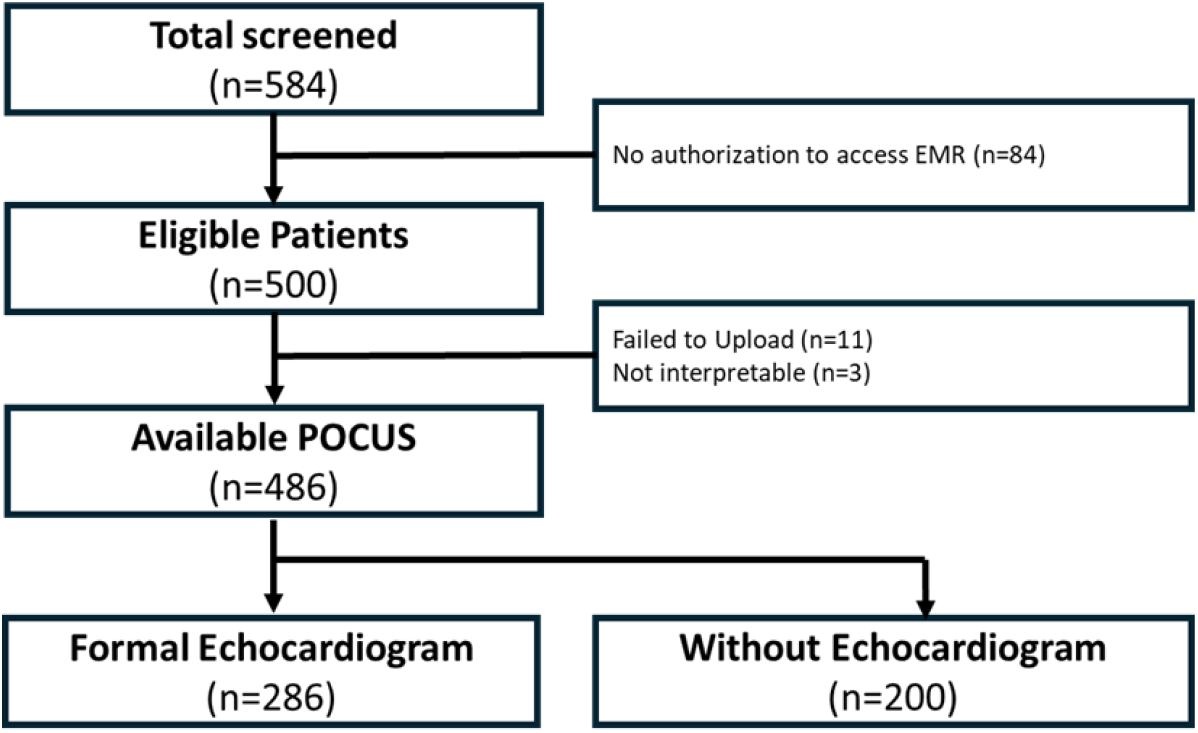
Study’s flowchart.

### Baseline characteristics

Patients had a median age of 64 years (IQR: 53.2-71.4) and 49% were female. Baseline characteristics of the study population across formal echocardiographic assessment availability status are provided in *Table 1*. Patients with available TTEs had higher rates of hypertension, diabetes, atrial fibrillation, and clinical heart failure but lower rates of coronary artery disease and chronic kidney disease. A total of ten patients had previous aortic valve replacement, all of whom had TTE evaluation. LVSD, HCM, and CA were diagnosed in 10%, 2%, and 2%, respectively. Of the patients with available formal TTE assessment, 50 (17.5%) had documented confirmed SHD.

**Table 1:** Baseline characteristics of the study population across formal TTE status.

TTE, transthoracic echocardiogram; S/P AVR, status post aortic valve replacement; HCM, hypertrophic
|  | Without TTE<br>(n=200) | With TTE<br>(n=286) | Total<br>(n=486) | p-value |
| --- | --- | --- | --- | --- |
| Age (median, IQR) | 61.0 (49.7,68.7) | 56.0 (57.0,73.9) | 64.0 (53.2, 71.4) | <0.01 |
| Female (%) | 112 (56) | 127 (44) | 239 (49) | 0.01 |
| Hypertension (%) | 50 (25) | 106 (37) | 156 (32) | <0.01 |
| Diabetes (%) | 17 (9) | 43 (15) | 60 (12) | 0.02 |
| Chronic Kidney Disease (%) | 14 (7) | 52 (18) | 66 (14) | <0.01 |
| Coronary artery disease (%) | 36 (18) | 97 (34) | 133 (27) | <0.01 |
| Heart failure (%) | 3 (1) | 75 (26) | 78 (16) | <0.01 |
| Valvular heart disease (%) | 0 (0) | 51 (18) | 51 (10) | <0.01 |
| AVR (%) | 0 (0) | 28 (10) | 28 (15) | <0.01 |
| Atrial fibrillation (%) | 9 (5) | 94 (33) | 103 (21) | <0.01 |
| HCM (%) | 0 (0) | 6 (2) | 6 (1%) | <0.01 |
| CA (%) | 0 (0) | 5 (2) | 5 (1%) | <0.01 |

cardiomyopathy; CA, cardiac amyloidosis.

### AI-ECG SHD classification

The median AI-ECG probability averages were 11.2% (IQR: 2.5-35.5) for AS, 0.96% (IQR: 0.41-3.1) for LVSD, 0.08% (IQR: 0.01-0.58) for (HCM), and 11.0% (IQR: 3.4-29.0) for CA. Most participants (67%) were negative for SHD by AI-ECG. Among those identified as “positive” for SHD, 21%, 8%, 3%, and 1% were positive for one, two, three, or four SHDs, respectively *Table 2*.

**Table 2:** AI-ECG SHD classification across the entire study population.

| Algorithm | AI ECG probability average (%) |  |  | p-value |
| --- | --- | --- | --- | --- |
|  | Without TTE (n=200) | With TTE<br>(n=286) | Total<br>(n=486) |  |
| AS | 6.0 (0.84-17.8) | 18.6 (0.8-17.8) | 11.2 (2.5-35.5) | <0.01 |
| LVSD | 0.62 (0.28-1.38) | 1.3 (0.51-5.8) | 0.96 (0.41-3.1) | <0.01 |
| HCM | 0.06 (0.01-0.25) | 0.12 (0.01-0.25) | 0.08 (0.01-0.58) | <0.01 |
| CA | 7.7 (1.9-17.6) | 14.7 (4.8-17.6) | 11.0 (3.4-29.0) | <0.01 |
| AI-ECG positive variables |  |  |  |  |
| 0 (%) | 163 (82) | 161 (56) | 324 (67) | <0.01 |
| 1 (%) | 31 (15) | 72 (25) | 103 (21) |  |
| 2 (%) | 5 (2) | 34 (12) | 39 (8) |  |
| 3 (%) | 1 (1) | 16 (6) | 17 (3) |  |
| 4 (%) | 0 (0) | 3 (1) | 3 (1) |  |

AI-ECG, artificial intelligence-enabled electrocardiogram; TTE, transthoracic echocardiogram; AS, aortic stenosis;
LVSD, left ventricular systolic dysfunction; HCM, hypertrophic cardiomyopathy; CA, cardiac amyloidosis.
Previously validated AI-ECG cutoffs are 40.6%, 25.6%, 11%, and 48.5% for AS, LVSD, HCM and CA, respectively.

Screening performance parameters using AI-ECG only are provided in *Figure 2*. Among the 286 patients with formal echocardiographic assessment, the NPV and PPV to detect any SHD by AI-ECG were 94% and 32%, respectively. The respective NPVs and PPVs for screening individual SHDs were 97% and 49% for LVSD, 89% and 38% for AS, and 99% and 15% for ILVWT (consistent with CA or HCM).

**Figure 2:**
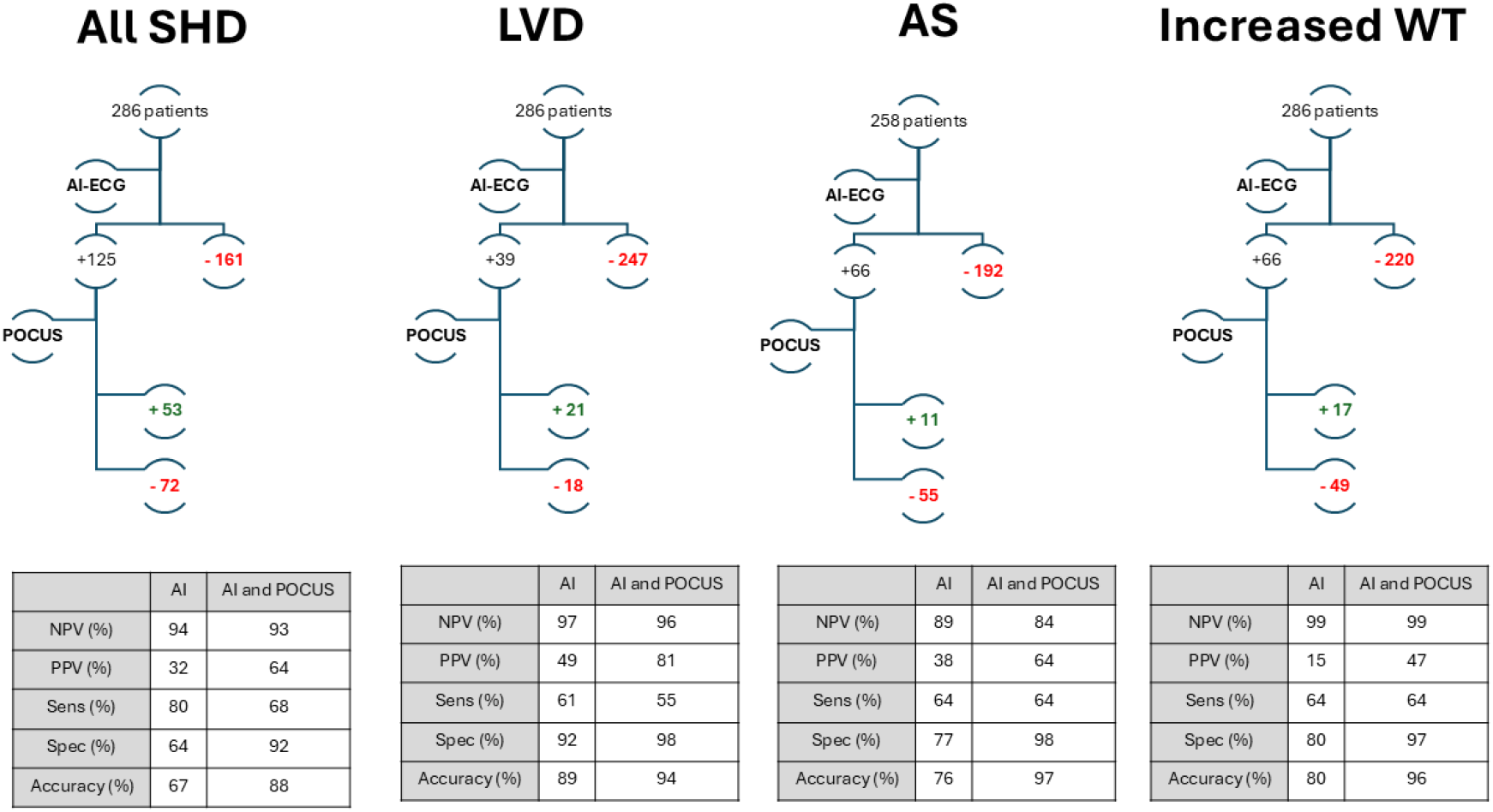
Predictive Value of Screening Using a POCUS-Integrated AI-ECG Approach.

### POCUS SHD classification

The predictive correlates utilizing the stepwise screening approach of AI-ECG followed by POCCUS are shown in *Figure 2*. Incorporating POCCUS as a second layer of screening for SHD in patients with formal TTE evaluation resulted in an increase in PPV to 64% and a slightly insignificant decrease in NPV to 93% to detect the existence of any SHD. The increase in PPV was consistent across all individual SHDs and was more prominent in less prevalent SHDs (AS). The NPVs were either unchanged or slightly, insignificantly, reduced across all examined SHDs. The respective NPVs and PPVs for screening individual SHDs were 96% and 81% for LVSD, 84% and 64% for AS, and 99% and 47% for ILVWT, consistent with CA or HCM.

Of the nineteen patients classified as “false-positives” by the integrated AI-ECG and POCCUS pathway, seventeen exhibited less than moderate structural heart disease (Graphical abstract): eight had aortic valve sclerosis or mild aortic stenosis, six showed ILVWT but without HCM or CA, two had normal ejection fraction with regional wall motion abnormalities, and one had severe right ventricular failure.

### Application of a two-layer screening method on a nonselective patient population

The application of a hypothetical screening method for SHD, first applying AI-ECG, followed by the application of POCCUS among AI-ECG “SHD-positive” patients, is presented in *Figure 3*. The number needed to screen, reflecting the number of patients necessary to undergo screening to identify one patient suspected of SHD eligible for formal TTE screening, was also calculated. When applying the hypothetical screening workflow on the entire patient population (n=486), the NNS to detect one patient with suspected SHD was 8. The NNS for individual SHDs was 22, 31, and 26 to detect one patient suspected to have LVSD, AS, or ILVWT consistent with CA or HCM.

**Figure 3:**
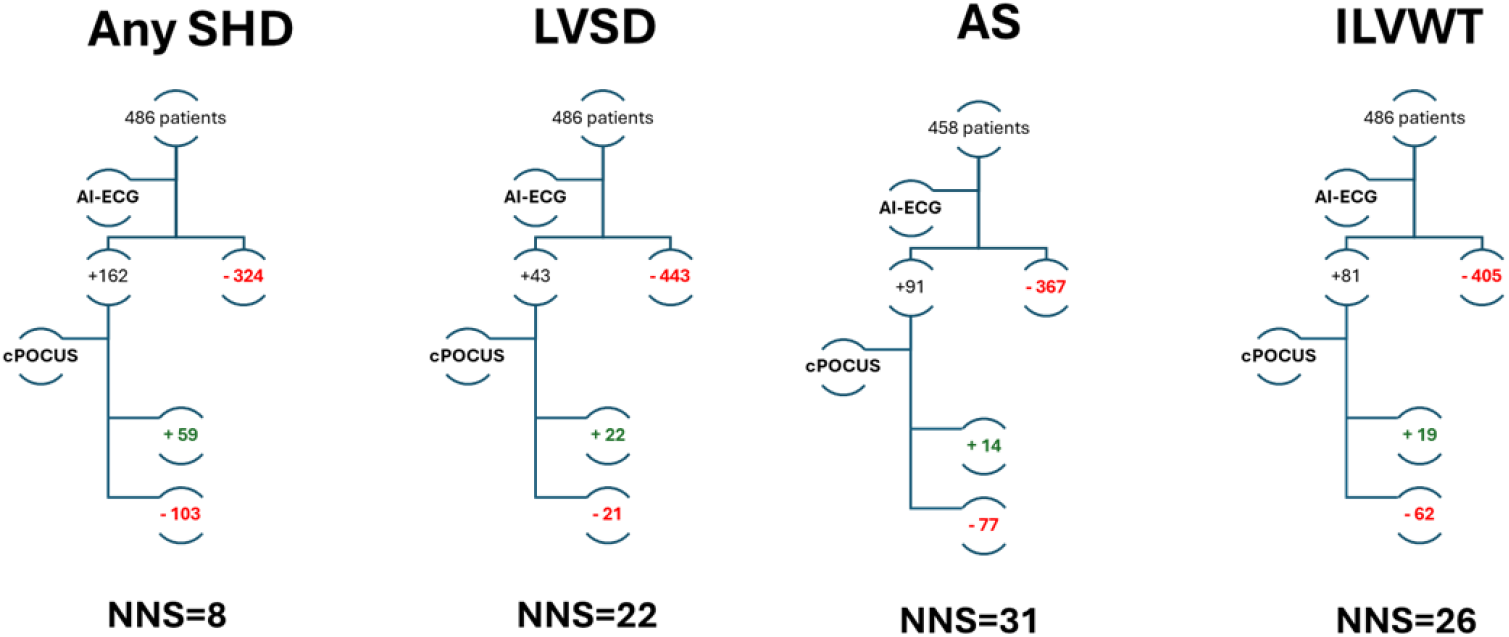
SHD screening with a POCUS-integrated AI-ECG approach in the entire cohort. AS, aortic stenosis; ILVWT, Increased Left Ventricle Wall Thickness; LVSD, left ventricular systolic dysfunction; SHD, Structural Heart Disease

## DISCUSSION

This study, conducted among 486 randomly selected outpatients referred for a clinically indicated ECG at the Mayo Clinic ECG laboratory, highlights the potential of AI-ECG and POCCUS for screening patients suspected of SHD. We evaluated a two-step screening approach: AI-ECG for initial triage of patients with low SHD probability, followed by POCCUS for AI-ECG-positive cases. Among 286 patients who underwent formal echocardiograms, this pathway demonstrated a 93% NPV and a 64% PPV, with higher values observed for specific SHD types such as LVEF and AS. Extrapolating across the entire study population, for screening purposes, the NNS was eight to identify one patient requiring formal echocardiographic evaluation. Importantly, among the nineteen patients classified as “false positives,” seventeen exhibited less-than-moderate or other coexisting SHD in their formal echocardiograms. This suggests that the two-step screening method had a PPV approaching 100% for detecting any type or severity of SHD. This study pioneers integrating AI-ECG and POCCUS in outpatient settings, offering a rapid, cost-effective, feasible, and standardized method for SHD screening (*Supplement 2*).

SHD represents a significant, potentially modifiable condition often characterized by asymptomatic or subclinical phases before manifesting symptomatic complications leading to morbidity or mortality. The four SHD types identified by current AI-ECG models carry substantial risks, including increased mortality rates during asymptomatic stages. However, implementing screening measures must consider factors such as disease prevalence, costs, feasibility, and patient benefit. While some SHDs, such as LVEF, are more prevalent in the general population, others, like AS, increase with age, and rare conditions, such as CA and HCM, pose specific challenges. Screening strategies typically require a clinical or epidemiological assessment of pretest probability. Given the lower likelihood of SHD in younger patients, effective screening might target those aged 50 years and older, balancing screening costs and clinical yield [13]. The study population’s mean age of 64 years and near-equal representation of both sexes make it a representative “target population” for such screening efforts. Of note, the disease prevalence observed in this proof-of-concept study differs from that reported in prior community-based trials (EAGLE), due to differences in study settings, as this study was conducted in an outpatient hospital environment rather than in the broader community, which may have influenced prevalence estimates. It is important to note, however, that the primary objective of the current study was to assess the feasibility and practicality of the proposed screening approach, rather than its large-scale effectiveness. The latter is currently being evaluated in an ongoing large-scale prospective clinical trial (NCT06891222). Furthermore, future screening strategies will likely be implemented in clinical or laboratory settings, particularly among patients referred for an ECG, rather than in community-based environments, where the application of POCCUS may present greater logistical challenges.

A previous randomized clinical trial investigating AI-ECG demonstrated a significant advancement in detecting LVSD (10). Additionally, a recent large-scale prospective echocardiographic screening study of asymptomatic individuals in the United Kingdom revealed a striking prevalence of 28.2% for any valvular heart disease (VHD), with 2.4% classified as moderate to severe (13). This finding aligns with the prevalence of 2.5% reported in the real-world setting study (14). Therefore, it is estimated that a substantial number of asymptomatic patients with VHD could be identified through proactive screening methods. Implementing an accessible, user-independent tool in community settings prior to formal echocardiography could potentially yield significant benefits in detecting clinically silent SHD.

Despite advancements in therapies, cost, and feasibility remain significant barriers. TTEs constitute a substantial portion of imaging expenses, often yielding no significant findings in primary care settings and having minimal impact on patient management in many cases [14,15]. Moreover, many asymptomatic community-dwelling patients are never referred for a TTE until presenting with symptoms or clinically evident disease. Limited TTE availability due to resource constraints in echocardiography laboratories complicates timely evaluation. Addressing these challenges, we advocate for integrating POCCUS following positive AI-ECG results to improve patient selection for formal echocardiographic assessment. Our two-tiered screening approach not only enhances PPV across all SHD types but also reveals that even false positives often uncover mild abnormalities warranting future monitoring. Notably,

POCCUS was performed by novice operators with minimal training, and image acquisition involved a continuous 10-second loop, increasing the likelihood of obtaining usable cardiac cycles. The evolution of AI and its expanded use in echocardiography [16,17] may eventually enable independent classification of POCCUS images based on segments of a single cardiac cycle or a fraction thereof.

Our study supports the widespread adoption of POCCUS-augmented AI-ECG as a screening tool, particularly in resource-limited settings lacking specialized expertise. Unlike other screening methods, such as mammography or colonoscopy, which demand extensive training, our study highlights that POCCUS can be effectively performed by novice operators after a brief two-day training period. Previous studies have demonstrated promising sensitivity and specificity for detecting abnormal LVEF using POCCUS, despite challenges such as suboptimal image quality [18,19]. In our study, adding POCCUS as a second layer of screening after AI-ECG led to a modest reduction in sensitivity, while significantly increasing specificity. However, it is crucial to highlight the distinct roles of each layer in our two-step screening approach. The first layer, AI-ECG, serves as a ‘screen-out’ layer, casting a wide net to exclude patients with a low probability of underlying SHD. The second layer, POCCUS, further refines the process by screening out AI-ECG-positive patients who are unlikely to have significant SHD and would not benefit from further evaluation with gold-standard diagnostic testing. While the addition of POCCUS introduces some modality-related limitations, its integration significantly increased specificity, despite a modest decrease in sensitivity, resulting in improved diagnostic accuracy, from 67% with AI-ECG alone to 88% with the two-layer screening.

This study has several limitations. Firstly, POCCUS, particularly when conducted by novice operators, may yield suboptimal imaging quality, potentially affecting interpretation. Additionally, limitations in the POCCUS review platform, including slow image uploads and measurement challenges, underscore the need for improved technological solutions. Despite these challenges, the study used trained echocardiographers to review and interpret POCCUS results, representing a proof-of-concept approach that could benefit from the future integration of dedicated AI algorithms tailored for POCCUS studies. Secondly, while randomly selected from outpatient settings undergoing an ECG, the study population may have a higher disease burden, as seen in the higher prevalence of disease, given clinical indications for ECG, limiting generalizability to broader screening populations. Thirdly, the study focused on four specific SHDs with validated AI-ECG algorithms using Mayo Clinic data that may not perform as well in other institutions. Future AI-ECG models could expand the workflow’s utility for other conditions, such as mitral regurgitation, tricuspid regurgitation, and constrictive pericarditis. Additionally, the recent development of an AI-ECG algorithm for determining a derived ECG phenotype common to most SHDs might offer an adjunct to screening patients with insufficient POCCUS imaging quality. However, its predictive value for specific SHDs remains to be confirmed in dedicated trials. Lastly, the current study offers a proof-of-concept platform

In conclusion, our study confirms the feasibility and accuracy of integrating POCCUS and AI-ECG for detecting SHD. This approach addresses current screening challenges and sets a precedent for future innovations in cardiovascular care. Looking forward, the incorporation of AI into POCCUS has the potential to revolutionize global SHD screening, offering scalable solutions comparable to recent advancements in large-scale echocardiogram analysis. Harnessing AI’s potential could reduce reliance on expert cardiologist reviews, expedite diagnostic timelines, and improve patient outcomes.

## Data Availability

All data referred to in this manuscript are available from the corresponding author upon reasonable request.

## AUTHOR CONTRIBUTIONS

SVP, GCK, and GT conceived and designed the study. FBA and GT analyzed the data and wrote the manuscript. FBA and RS acquired and collected the data. EL provided technical support. JB, AJB, VTN, JKO, PAN, PAP, AZ FLJ, and PAF provided essential input on data analysis approaches and reviewed the manuscript.

## FIGURE LEGENDS

**Central Figure: Detecting Structural Heart Disease with AI-ECG and POCCUS integration**

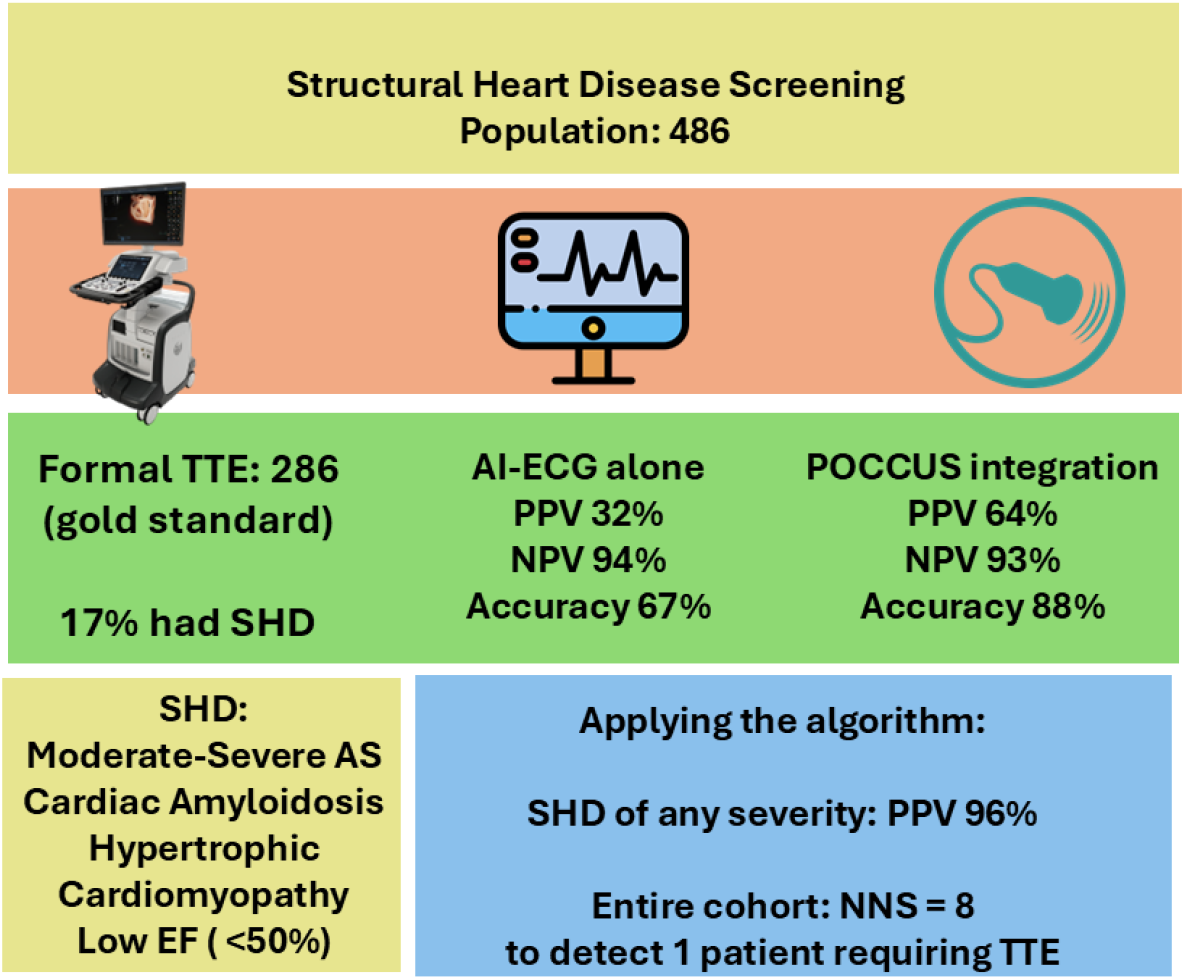

